# Exploring the adoption of digital pathology in clinical settings - Insights from a cross-continent study

**DOI:** 10.1101/2023.04.03.23288066

**Authors:** Daniel Gomes Pinto, Andrey Bychkov, Naoko Tsuyama, Junya Fukuoka, Catarina Eloy

**Author notes:** **Corresponding Author:** Catarina Eloy, M.D., Ph.D., IPATIMUP, Rua Júlio Amaral de Carvalho 45, Porto, 4200-135, Portugal. author mail.

## Abstract

The last seventy years have been characterized by rapid advancements in computer technology, and the healthcare system has not been immune to this trend. However, anatomic pathology has remained largely an analog discipline. In recent years, this has been changing with the growing adoption of digital pathology, partly driven by the potential of computer-aided diagnosis. As part of an international collaboration, we conducted a comprehensive survey to gain a deeper understanding of the status of digital pathology implementation in Europe and Asia. A total of 127 anatomic pathology laboratories participated in the survey, including 75 from Europe and 52 from Asia, with 72 laboratories having established digital pathology workflow and 55 without digital pathology. Laboratories using digital pathology were thoroughly questioned about their implementation strategies and institutional experiences, including details on equipment, storage, integration with laboratory information system, computer-aided diagnosis, and the costs of going digital. The impact of the digital pathology workflow was also evaluated, focusing on turnaround time, specimen traceability, quality control, and overall satisfaction. Laboratories without access to digital pathology were asked to provide insights into their perceptions of the technology, expectations, barriers to adoption, and potential facilitators. Our findings indicate that while digital pathology is still the future for many, it is already the present for some. This decade may be a time when anatomic pathology finally embraces the digital revolution on a large scale.

**HIGHLIGHTS:**

- Larger labs adopt digital pathology more
- Full digital transition is still rare nowadays
- Many initial concerns have not materialized after implementation
- Most non-digital laboratories plan to go digital soon

## INTRODUCTION

The mid-to-late 20th century and the early 21st century were significantly shaped by advancements in computer technology, and healthcare systems were not exempt from these developments^1^. The pace of technological adoption varied among medical specialties, with radiology being a prime example of a field that embraced technology quickly. Film was replaced by digital image sensors for radiological use in the 1980s, and by the late 1990s, most radiology departments in developed countries had fully transitioned to digital^2^. On the other hand, anatomic pathology has largely remained an analog discipline. Histopathology and cytopathology practices also rely on image interpretation, but a digital sensor for the direct capture of histological images from unprocessed tissue has yet to prove viable in routine practice. This means that anatomical pathology continues to use traditional techniques for glass slide preparation, staining, and bright- field microscope observation^3^. Efforts to incorporate digital technologies into clinical practice in anatomic pathology began in the late 1980s, but these were only used in niche contexts and never became mainstream^4^. The conversion of glass slides into digital whole-slide images (WSIs) resulted in increased processing times and impractical storage requirements, which, being combined with the high costs of implementation, insufficient display quality, and regulatory hurdles, prevented widespread adoption of the technology^5,6^.

The landscape of digital pathology has changed in recent years. Display and storage technologies have improved and become more accessible, while processing power and slide scanner technology have advanced dramatically. These devices, which are essentially image sensors coupled to microscopes with varying degrees of automation, have become more sophisticated, reducing scanning times and costs. Furthermore, recent developments in artificial intelligence (AI) research have brought the promise of a revolution in pathology, making digital pathology more attractive^5,7,8^. This has led to a significantly increased adoption of digital workflows, with multiple institutions publishing their experiences over the past decade^9-20^. Several national and international associations have also released guidelines for the adoption and implementation of digital pathology^7,21-23^.

Surveys on the topic have been developed recently, aiming at better understanding the perceptions of pathologists regarding digital pathology and its impacts on routine practice^20,24-30^. These surveys focused on the implementation status in individual countries such as the United Kingdom, Switzerland, Canada, India, and others, but none of them went beyond a national level.

To gain a deeper understanding of the status of digital pathology implementation in two major markets in the world – Europe and Asia, we conducted a comprehensive survey. In this publication, we present the results of the survey and discuss the most important findings.

## MATERIAL AND METHODS

Over the course of several virtual meetings, the authors collaborated to draft a comprehensive survey that covers various topics deemed relevant based on expert opinion and current literature. The survey was created using the Google Forms (Google LLC, Mountain View, CA, USA) platform and was distributed to a small group of ten participants for pilot testing. After considering the feedback and regional differences, the survey was finalized and sent directly to heads of pathology departments in Europe and Asia. The mailing list was created based on relevant publications on digital pathology and personal contacts of the authors. Only one participant was allowed per institution.

The survey consisted of several sections, using different types of answer options, such as multiple-choice questions with a single or multiple responses, Likert scales (from 1 to 5), and short open-ended questions. The participants were asked to provide general information about themselves and their institution and whether the workflow in their laboratory relied on digital pathology.

If the institution had no digital pathology workflow, the respondents were directed to a standalone section focused on their experience with WSIs outside of routine practice, future vision for the implementation of digital pathology, perception of digital pathology, and possible drivers and hindrances of adoption. Alternatively, if the institution had any use of digital pathology workflow (with an operating slide scanner as a prerequisite), the respondents were directed to an in-depth survey covering the following sections: 1) implementation, 2) equipment and storage, 3) software, and 4) the impact of digital pathology on laboratory routine. A flowchart illustrating the structure of the survey can be seen in Figure 1. The full list of survey questions is provided in the Supplementary Table 1.

**Figure 1:**
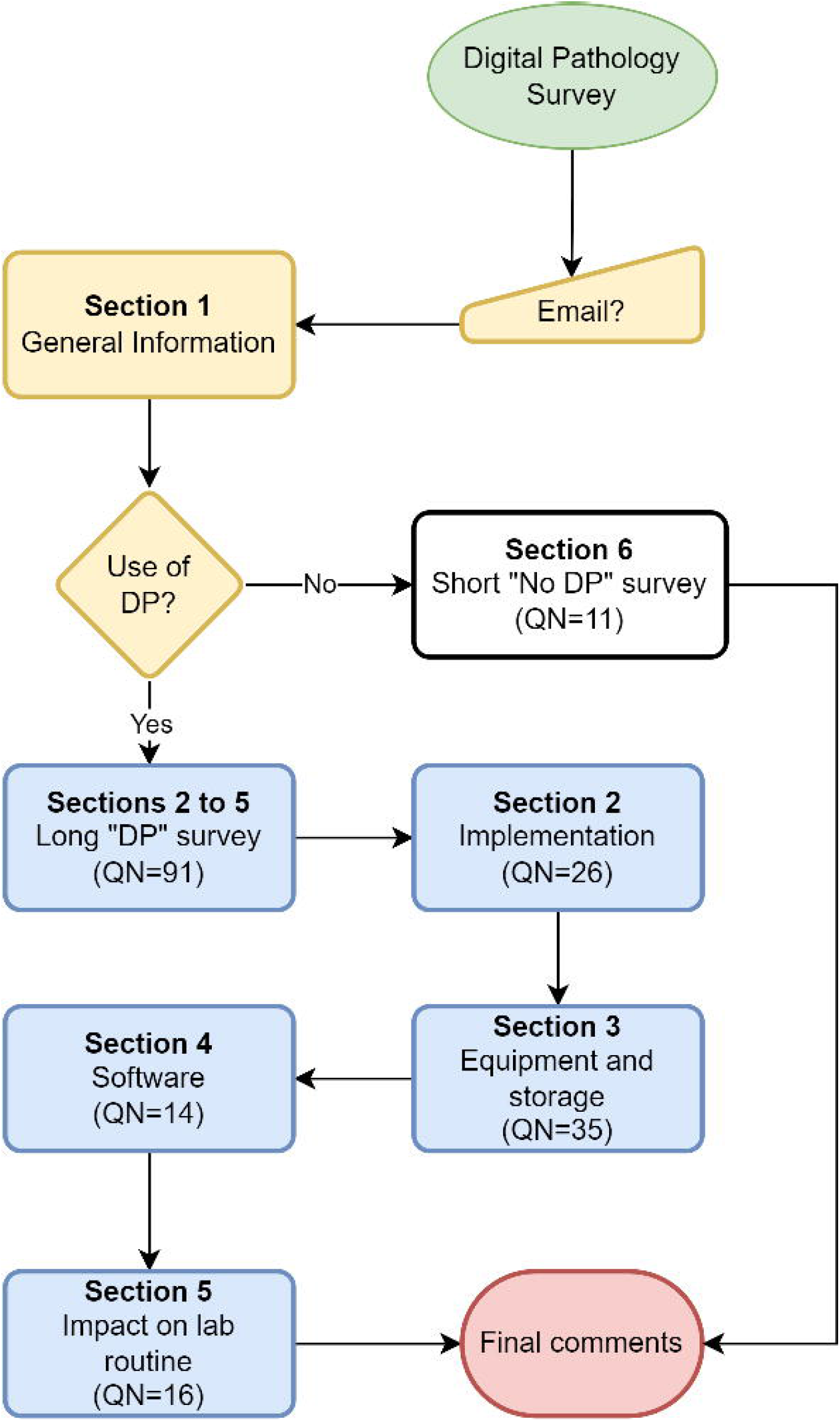
Flowchart of the survey DP, digital pathology; QN, number of questions

The survey was open for twenty weeks from July 20 to December 13, 2020, and the responses were collected in a Microsoft Excel spreadsheet (Microsoft, Redmond, WA, USA). After the closing date, the database was screened for duplicate, incomplete, or invalid responses, which were excluded, and the remaining valid responses were analyzed. Descriptive analysis was performed using Microsoft Excel, and statistical analysis was performed using GraphPad Prism 9 (GraphPad Software Inc., San Diego, CA, USA). Nominal variables were described in terms of frequencies, and ordinal and continuous variables were described using means and medians. Statistical significance was tested using chi-squared test or Fisher’s exact test in nominal variable, and Student’s t-test or Mann-Whitney U test in ordinal and continuous variables. The results were considered statistically significant if the p-value was less than 0.05.

## RESULTS

A total of 127 responses were collected, with 72 coming from laboratories that have fully or partially implemented a digital pathology workflow and 55 from laboratories without in-house digital pathology. Continent-wise, this study enrolled 75 European and 52 Asian institutions. The statistical analysis indicated that out of the 129 survey questions only 7 showed statistically significant differences between the European and Asian cohort (Supplementary Table 2–5). These differences were found in the sections addressed to laboratories with a digital pathology workflow. Therefore, the results of the survey are combined from both regions, unless specified otherwise.

Seventy-two laboratories with a digital pathology workflow participated in the survey, with roughly equal representation from Asia (n = 38; 52.8%) and Europe (n = 34; 47.2%). These digital adopters laboratories were from 14 European and 7 Asian countries. The majority of respondents were from university hospitals with moderate to high annual caseloads, followed by cancer centers, while private laboratories made up a minority. A summary of the geographic distribution and caseload of laboratories can be found in Figure 2. Digital pathology laboratories were usually staffed with 6 to 10 (n = 23; 31.9%), 1 to 5 (n = 17; 23.6%), 11 to 20 (n = 17; 23.6%), or 21 to 40 (n = 13; 18.1%) full-time pathologists. While some labs had no part-time pathologists (n = 13; 18.1%), most had at least one to five part-time pathologists (n = 49; 68.1%) on staff to help manage the workload and provide expertise in specific areas of pathology. Most of the institutions that adopted digital pathology were involved in undergraduate medical education (72.2%) and postgraduate training in anatomic pathology (83.3%).

**Figure 2:**
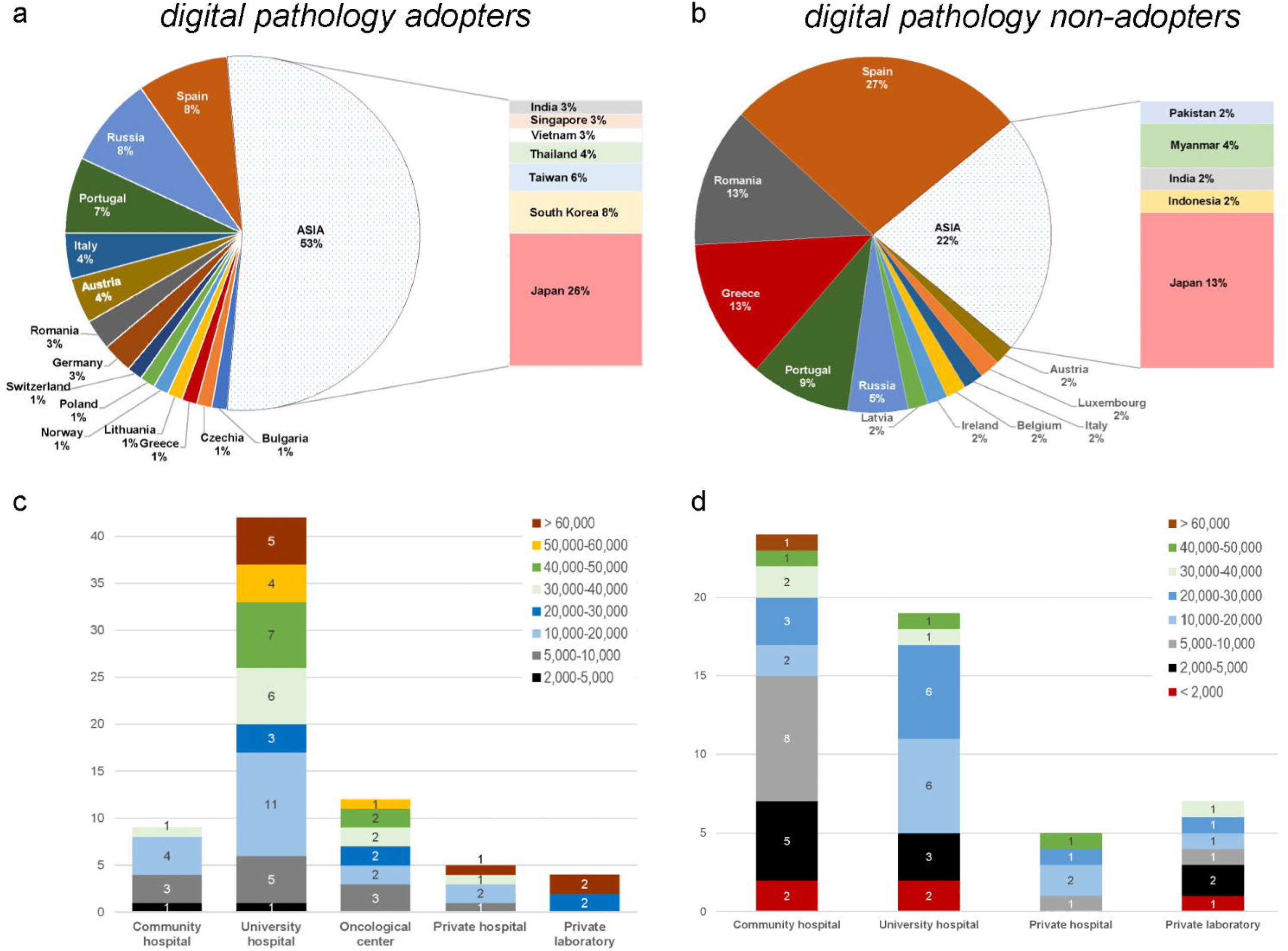
General information about participants Distribution per country and caseload per institution type among adopters (**A** and **C**, respectively) and non-adopters (**B** and **D**, respectively) of digital pathology.

Regarding the overall use of digital pathology, most of the respondents reported that they only digitize cases for specific purposes, such as teaching and research (n = 43; 59.7%). This was followed by those who digitize most of their workload (n = 17; 23.7%), and those who routinely digitize only selected pathology subspecialties, such as gastrointestinal pathology, breast pathology, or dermatopathology (n = 12; 16.7%).

### Implementation

In the cohort of digital pathology adopters, implementation mainly took place from 2015 onwards (n = 49; 68.1%), with notable exceptions in both Europe and Asia, where single laboratories started digitizing slides in 2003 and 2004, respectively. In terms of strategy, the preferred approaches were to start by digitizing biopsies (n = 15; 20.8%) and/or cases from select subspecialities (n = 22; 30.6%). Furthermore, most institutions digitized cases for educational purposes and/or for discussion at tumor boards (n = 43; 59.7%). Of note, only twelve (16.7%) institutions immediately started digitizing all their cases after installation of the slide scanner.

Regarding current uses, the majority of the respondents stated that they digitize most specimen types and ancillary tests, including biopsies, surgical specimens, and immunohistochemistry (IHC) slides (n = 68; 94.4%, n = 57; 79.2% and n = 58; 80.6%, respectively). Twenty-six participants (36.1%) indicated that they rely on digital pathology to enable the use of computer-aided-diagnosis (CAD) algorithms. Cytology is only digitized by a few laboratories (n = 6; 18.2%), but other than that, there was no clear preference for the subspecialty digitized, with an average of 50% laboratories digitizing all areas either partially or fully, with partial scanning being the most common option. In terms of caseload, half of the respondents digitized less than 5% of the total volume (n = 36; 50%); however, 16 institutions (22.2%) scan more than 75% of their caseload. From these, 14 (19.4%) laboratories currently digitize all their histology routine, including IHC, and another 15 (20.8%) expected to reach this point within 2 years (n = 15; 20.8%). Interestingly, despite the low percentage of total cases digitized in many laboratories, almost all institutions stated that they run their scanners either every working day, or at least 2 to 3 days per week (n = 60; 83.3%). Approaches to scanning varied, with an even split on digitizing cases before or after diagnosis, with some respondents utilizing both approaches. Retrospective scanning, mainly for research purposes, is performed by most institutions (n = 55; 76.4%).

Pre-scanning quality control is routinely performed either by technicians or pathologists in a similar proportion (n = 35; 48.6% vs. n = 34; 47.2%); however, only a minority of laboratories records the scanning error rate (n = 21; 29.2%). Among the latter, a low error rate ≤ 1% was documented (n = 11; 52.4%). Validation protocols were adopted by less than half institutions (n = 41; 56.9%), mainly based on the recommendations by the College of American Pathologists^23^ (n = 15; 20.8%) or those from national societies (n = 11; 15.3%). Sample traceability is an important part of quality control, allowing for better and directed troubleshooting^7^. Accordingly, most laboratories equipped with slide scanners employed 1D/2D barcoded slides (n = 60; 83.3%) and cassettes (n = 44; 61.1%). Photography in the grossing room (n = 35; 48.6%) and barcoded containers in the wet archive (n = 39; 54.2%) were also commonly in use for sample traceability, while videorecording remained a relatively niche tool, with only 7 (9.7%) of respondents mentioning its usage.

Traditional microscopes are still used in most of the digitized laboratories, including those that have undergone a fully digital transition, mainly for the purpose of viewing slides under polarizing light, cytology slides, and microorganism stains (n = 56; 77.8%, n = 49; 68.1%, and n = 38; 52.8%, respectively). The most cited reasons for favoring traditional microscopes in these cases were the lack of the required scanner mode, such as Z-stacking for cytology (n = 17; 51.5%), and insufficient WSI quality (n = 32; 44.4%). Slightly more than a third of the laboratories digitized frozen sections (n = 28; 39%).

In terms of use by pathologists, some institutions had all staff pathologists using WSIs for diagnostic work (n = 15, 20.8%), while the majority had lower digital penetrance, with usage as low as 25% (n = 41, 56.9%). Personal opposition to digital pathology was cited as the main reason for low adherence (n = 30, 41.7%), followed by concerns over a slower turnaround time for digitized cases (n = 23, 31.9%). Statistically significant differences were found between the continents, with Asian pathologists more likely to show prejudice against digital pathology (n = 20, 51.3% vs. n = 10, 30.3%), have limited access to digital pathology workstations (n = 12, 30.8% vs. n = 6, 18.2%), and be more concerned about a slower turnaround time (n = 19, 48.7% vs. n = 4, 12.1%). Supplementary Table 4 summarizes the questions that showed statistically significant differences between the European and Asian cohorts of participants.

The implementation of a digital workflow requires modifications to the usual laboratory processes to some extent^7^. These adaptations, mainly involving tasks performed by laboratory technicians, included performing thinner sections for easier focusing (n = 25, 34.7%), placing two sections of the same biopsy on a single slide to minimize rescan needs (n = 17, 23.6%), placing sections closer together and in the direction of scanning to achieve faster scanning times (n = 15, 20.8%), and dividing large specimens to avoid potential focusing issues (n = 14, 19.4%). Physical space adaptations were also deemed necessary, with most respondents requiring changes with or without a need for an actual space expansion (n = 49, 58.1% and n = 9, 12.5%, respectively).

While technological advances have facilitated wider implementation of digital pathology, challenges persist in its daily use. Respondents reported difficulty in recognizing fine details on digital slides, such as microorganisms (n = 38; 52.8%). A significant portion also complained about slow image loading speed on viewing devices (n = 38; 52.8%), presence of out-of-focus areas (n = 36; 50%), and lack of functionality comparable to manual fine focusing on traditional microscopes (n = 32; 44.4%). A few respondents also reported nuclear chromatic aberrations (n = 7; 9.7%). However, the same number of respondents reported no problems with digital slides.

The adoption of new technologies and workflows often necessitates the hiring of new employees. In this regard, significant geographic differences were found, with a majority of digital pathology adopters in Europe claiming no new staff was hired, while this was not the case in Asia (n = 26; 78.8% vs. n = 15; 38.5%, respectively; p < 0.01). Technicians were the most commonly recruited staff position in both regions (n = 5; 15.2% vs. n = 17; 43.6%, respectively) to support digital transformation. In Asia, secretaries were also frequently hired, unlike in Europe (n = 11; 28.2% vs. n = 1; 3.0%, respectively).

Training for existing staff is also often necessary and, in this series, it was mostly done through on-site training by industry representatives (n = 47; 65.3%), followed by centralized industry seminars and conferences (n = 30; 41.7%, each). However, the sufficiency of such training is debatable, with an almost even split between “yes” and “no” responses (n = 36; 50.0% vs. n = 35; 48.6%).

### Equipment

The centerpieces of any modern laboratory equipped with digital pathology are slide scanners. In our series of digital pathology adopters, most institutions had either one or two scanners (76.4% and 23.6% respectively, with a total of 55 respondents). The median pooled capacity was 210.0 slides per institution, with an interquartile range (IQR) of 541.0. Leica/Aperio was the most popular brand across all scanner types, i.e., high-, medium-, and low-throughput, see details in Table 2. Only a few respondents (18%) reported using single slide scanners. A majority of participants (66.7%) mentioned that in-vitro diagnostics (IVD) certification was a priority when purchasing a scanner. In terms of advanced features, such as Z-stacking, extended focus mode (also known as extended depth of field), or dark-field microscopy, most respondents said that the available scanner either does not support them or, even if it does, the pathologists do not use them for clinical purposes (n = 65; 90.2%, n = 43; 59.7%, and n = 60; 89.3%, for each feature, respectively). In terms of magnification, most respondents preferred 40× scanning protocols, but believed that 20× was acceptable for most cases (n = 39; 54.2%). Those who stated that 20× scanning was sufficient for an accurate diagnosis were in a similar proportion to those who argued for digitizing exclusively at 40× (n = 14; 19.4% vs. n = 13; 18.1%).

**Table 1:**
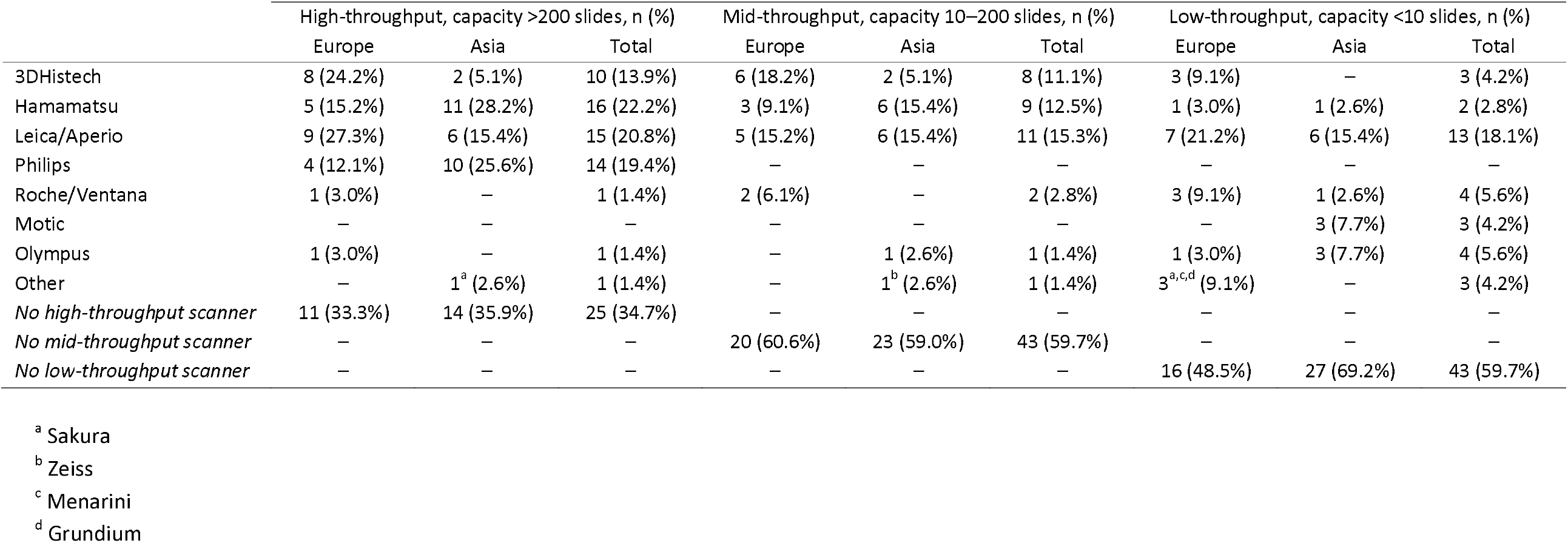
Scanner brands

**Table 2:** Monitors used for digital pathology

|  | Europe, n (%) | Asia, n (%) | Total, n (%) |
| --- | --- | --- | --- |
| Acer | 1 (3.0%) | – | 1 (1.4%) |
| Apple | 1 (3.0%) | 2 (5.1%) | 3 (4.2%) |
| Asus | 1 (3.0%) | – | 1 (1.4%) |
| Barco<sup>a</sup> | 1 (3.0%) | 1 (3.0%) | 2 (2.8%) |
| BENQ | – | 1 (2.6%) | 1 (1.4%) |
| Dell | 6 (18.2%) | 7 (17.9%) | 13 (18.1%) |
| EIZO<sup>b</sup> | 1 (3.0%) | 6 (15.4%) | 7 (9.7%) |
| Fujitsu | 1 (3.0%) | – | 1 (1.4%) |
| HP | 9 (27.3%) | 3 (7.7%) | 11 (15.3%) |
| Iiyama | 1 (3.0%) | – | 1 (1.4%) |
| JVC | – | 1 (2.6%) | 1 (1.4%) |
| Lenovo | 1 (3.0%) | – | 1 (1.4%) |
| LG | 2 (6.1%) | 2 (5.1%) | 4 (5.6%) |
| NEC | – | 1 (2.6%) | 1 (1.4%) |
| Olympus<sup>a</sup> | – | 1 (2.6%) | 1 (1.4%) |
| Samsung | 1 (3.0%) | 2 (5.1%) | 3 (4.2%) |
| Philips | – | 1 (2.6%) | 1 (1.4%) |
| Sharp | 1 (3.0%) | – | 1 (1.4%) |

<sup>a</sup> medical grade<sup>b</sup> professional grade

A pathologist’s workstation, typically consisting of a personal computer and a monitor, is another key component of a digital pathology suite. The median number of workstations per institution was 2.0 (IQR, 9.0; range, 1–45), with a median of 2.0 screens per workstation (IQR, 1.0; range, 1–3). The median screen size used for digital pathology was 27.0 inches (IQR, 4.5; range, 17–51 inches). There was a statistically significant difference between European and Asian institutions in this regard, with European screens being larger by approximately two inches (median 27.0 inches, range 21–51 vs. median 25.0 inches, range 17–48). The most prevalent screen resolution was 1080p (1920 × 1080 progressively displayed pixels, or full HD), chosen by 37.5% of respondents (n = 27). The second most common resolution was 4K (n = 17, 23.6%), followed by 2K (n = 10, 13.9%), while a minority used resolutions lower than 1080p (n = 5, 6.9%). A considerable proportion of respondents were not aware of their screen resolution (n = 10, 13.9%).

Most of the screens used for digital pathology were “consumer-grade,” meaning they are marketed to regular consumers (n = 47; 65.3%). At the same time, professional and medical-grade screens were also used in more than half of the institutions (n = 39; 54.2%). The majority of screens were based on liquid crystal display (LCD) panels, with a preponderance of in-plane switching (IPS) panels (n = 22; 30.6% and n = 17; 23.6%, respectively). The use of organic light emitting diode (OLED) panels was very limited (n = 5; 6.9%). A list of monitor brands used for digital pathology is provided in Table 3. The proportion of laboratories that color-calibrated their screens was equal to those that did not (n = 23; 31.9%). Most institutions (n = 42; 58.3%) did not place restrictions on the use of smaller screens, such as those of smartphones, tablets, or laptops, for digital pathology. However, a significant proportion of respondents discouraged their use for diagnostic purposes, with only a few having outright prohibitions (n = 26; 36.1% vs. n = 4; 5.6%).

**Table 3:** Subjective perception of digital pathology implementation

|  | Strongly disagree, n (%) |  |  | Disagree, n (%) |  |  | Neither agree nor disagree, n (%) |  |  | Agree, n (%) |  |  | Strongly agree, n (%) |  |  |
| --- | --- | --- | --- | --- | --- | --- | --- | --- | --- | --- | --- | --- | --- | --- | --- |
|  | Europe | Asia | Total | Europe | Asia | Total | Europe | Asia | Total | Europe | Asia | Total | Europe | Asia | Total |
| Improved turnaround time | 7 (21.2%) | 5 (12.8%) | 12 (16.7%) | 8 (24.2%) | 11 (28.2%) | 19 (26.4%) | 10 (30.3%) | 14 (35.9%) | 24 (33.3%) | 6 (18.2%) | 8 (20.5%) | 14 (19.4%) | 2 (6.1%) | 1 (2.6%) | 3 (4.2%) |
| Improved sample traceability | 5 (15.2%) | 2 (5.1%) | 7 (9.7%) | 3 (9.1%) | 2 (5.1%) | 5 (6.9%) | 10 (30.3%) | 12 (30.8%) | 22 (30.6%) | 5 (15.2%) | 12 (30.8%) | 17 (23.6%) | 10 (30.3%) | 11 (28.2%) | 21 (29.2%) |
| Improved collaboration between peers in the lab | 3 (9.1%) | 1 (2.6%) | 4 (5.6%) | 1 (3.0%) | 2 (5.1%) | 3 (4.2%) | 13 (39.4%) | 11 (28.2%) | 24 (33.3%) | 9 (27.3%) | 15 (38.5%) | 24 (33.3%) | 7 (21.2%) | 10 (25.6%) | 17 (23.6%) |
| Improved the response time of consultation cases | 4 (12.1%) | 1 (2.6%) | 5 (6.9%) | 4 (12.1%) | 2 (5.1%) | 6 (8.3%) | 12 (36.4%) | 10 (25.6%) | 22 (30.6%) | 6 (18.2%) | 11 (28.2%) | 17 (23.6%) | 7 (21.2%) | 15 (38.5%) | 22 (30.6%) |
| Useful for teaching | 1 (3.0%) | 0 | 1 (1.4%) | 0 (0.0%) | 0 (0.0%) | 0 (0.0%) | 1 (3.0%) | 3 (7.7%) | 4 (5.6%) | 8 (24.2%) | 12 (30.8%) | 20 (27.8%) | 23 (69.7%) | 24 (61.5%) | 47 (65.3%) |
| Useful in interdisciplinary meetings | 4 (12.1%) | 1 (2.6%) | 5 (6.9%) | 4 (12.1%) | 2 (5.1%) | 6 (8.3%) | 12 (36.4%) | 10 (25.6%) | 22 (30.6%) | 6 (18.2%) | 11 (28.2%) | 17 (23.6%) | 7 (21.2%) | 15 (38.5%) | 22 (30.6%) |
| Led to a decrease in the use of paper | 2 (6.1%) | 0 | 2 (2.8%) | 1 (3.0%) | 0 (0.0%) | 1 (1.4%) | 3 (9.1%) | 2 (5.1%) | 5 (6.9%) | 8 (24.2%) | 10 (25.6%) | 18 (25.0%) | 19 (57.6%) | 27 (69.2%) | 46 (63.9%) |
| Required technical improvements in slides preparation | 7 (21.2%) | 3 (7.7%) | 10 (13.9%) | 3 (9.1%) | 6 (15.4%) | 9 (12.5%) | 10 (30.3%) | 11 (28.2%) | 21 (29.2%) | 3 (9.1%) | 12 (30.8%) | 15 (20.8%) | 10 (30.3%) | 7 (17.9%) | 17 (23.6%) |
| Improved the technical quality of slides in the lab | 3 (9.1%) | 0 | 3 (4.2%) | 3 (9.1%) | 4 (10.3%) | 7 (9.7%) | 9 (27.3%) | 8 (20.5%) | 17 (23.6%) | 10 (30.3%) | 18 (46.2%) | 28 (38.9%) | 8 (24.2%) | 9 (23.1%) | 17 (23.6%) |
| Clinicians are aware of it | 3 (9.1%) | 0 | 3 (4.2%) | 3 (9.1%) | 6 (15.4%) | 9 (12.5%) | 14 (42.4%) | 18 (46.2%) | 32 (44.4%) | 9 (27.3%) | 11 (28.2%) | 20 (27.8%) | 4 (12.1%) | 4 (10.3%) | 22 (30.6%) |
| Clinicians are aware of its capabilities | 6 (18.2%) | 3 (7.7%) | 9 (12.5%) | 4 (12.1%) | 8 (20.5%) | 12 (16.7%) | 9 (27.3%) | 5 (12.8%) | 14 (19.4%) | 10 (30.3%) | 16 (41.0%) | 26 (36.1%) | 4 (12.1%) | 7 (17.9%) | 11 (15.3%) |
| Improved diagnostic accuracy | 11 (33.3%) | 4 (10.3%) | 15 (20.8%) | 2 (6.1%) | 12 (30.8%) | 14 (19.4%) | 10 (30.3%) | 10 (25.6%) | 20 (27.8%) | 9 (27.3%) | 8 (20.5%) | 17 (23.6%) | 1 (3.0%) | 5 (12.8%) | 6 (8.3%) |
| Lead to better quality control | 5 (15.2%) | 3 (7.7%) | 8 (11.1%) | 5 (15.2%) | 9 (23.1%) | 14 (19.4%) | 12 (36.4%) | 17 (43.6%) | 29 (40.3%) | 7 (21.2%) | 9 (23.1%) | 16 (22.2%) | 4 (12.1%) | 1 (2.6%) | 5 (6.9%) |
| Allowed my lab to increase its caseload | 4 (12.1%) | 2 (5.1%) | 6 (8.3%) | 3 (9.1%) | 6 (15.4%) | 9 (12.5%) | 8 (24.2%) | 11 (28.2%) | 19 (26.4%) | 12 (36.4%) | 16 (41.0%) | 28 (38.9%) | 6 (18.2%) | 4 (10.3%) | 10 (13.9%) |
| I would step back to analog microscopy If I could | 11 (33.3%) | 5 (12.8%) | 16 (22.2%) | 7 (21.2%) | 6 (15.4%) | 13 (18.1%) | 9 (27.3%) | 17 (43.6%) | 26 (36.1%) | 2 (6.1%) | 10 (25.6%) | 12 (16.7%) | 4 (12.1%) | 1 (2.6%) | 5 (6.9%) |
| Its value is understood by the hospital administration | 23 (69.7%) | 17 (43.6%) | 40 (55.6%) | 4 (12.1%) | 10 (25.6%) | 14 (19.4%) | 6 (18.2%) | 8 (20.5%) | 14 (19.4%) | 0 (0.0%) | 1 (2.6%) | 1 (1.4%) | 0 | 3 (7.7%) | 3 (4.2%) |

Robotic microscopes, which enable real-time telepathology for consultation or frozen section cases or even the creation of WSIs, are also worth mentioning. A large majority of respondents did not have access to these devices (n = 65; 90.3%), while only a few still used them for diagnostic purposes (n = 3; 4.2%). Only 5 institutions claimed to use a robotic microscope with remote navigation for live-stream microscopy (n = 5; 6.9%).

### Storage

WSIs consume vast amounts of digital memory, therefore all labs implementing a digital workflow must carefully consider their storage configuration and policies before installing a scanner. Regulations on the required retention period for WSIs vary by country, if regulated at all. In our series, most laboratories with slide scanners stored WSIs for a year or longer (n = 49; 68.1%), with only a small minority of 4 institutions (5.6%, all from Europe) discarding them within a few months. European pathologists were less likely to store slides for a year or longer compared to their Asian counterparts (n = 17; 51.5% vs. 32; 82.1%) and more likely to store virtual slides for a short period of a year or less (n = 7; 21.2% vs. n = 1; 2.6%). These differences were statistically significant (p < 0.01).

Most respondents consider storage planning to be important, stating that future storage needs were calculated at the time of implementation and that they estimated the capacity needed in the mid-to-long term (n = 44; 61.1% each). Based on the caseload, 30 institutions (41.7%) reported a requirement to add 5 terabytes (TB) or less of data to their archive every year, while 8 respondents (11.1%) added more than 50 TB of data to their archive each year. This storage was mostly not tiered (n = 38; 52.8%), with more than quarter opting for additional cold storage (n = 19; 26.4%). Data was stored on remote servers accessed over the internet (“cloud storage”) by a significant number of respondents (n = 23; 31.9%), enabled by high internet speed. The median download speed was 106.0 (IQR, 410.0; range, 8–10000) megabits per second (mbps), and the median upload speed was 97.5 (IQR, 176.0; range, 10–750) mbps.

## Funding

Funding for laboratory equipment is a sensitive topic, and a substantial number of respondents chose not to disclose their institution’s expenses for digital pathology implementation (n = 26; 36.1%). Of those that did share, most spent USD 100,000–1,000,000 (n = 29; 63%), ten institutions (21.7%) spent between USD 1,000,000 and 5,000,000 and one (2.2%) spent more than USD 5,000,000. Most institutions funded their digital transition using own funds (n = 50; 69.4%), followed by regional or national level projects. Most respondents expected a return on their investment either in the medium or long term (n = 26; 36.1%), while approximately one-fifth admitted they did not expect any returns (n = 15; 20.8%). A detailed representation of the funding data can be seen in Figure 3.

**Figure 3:**
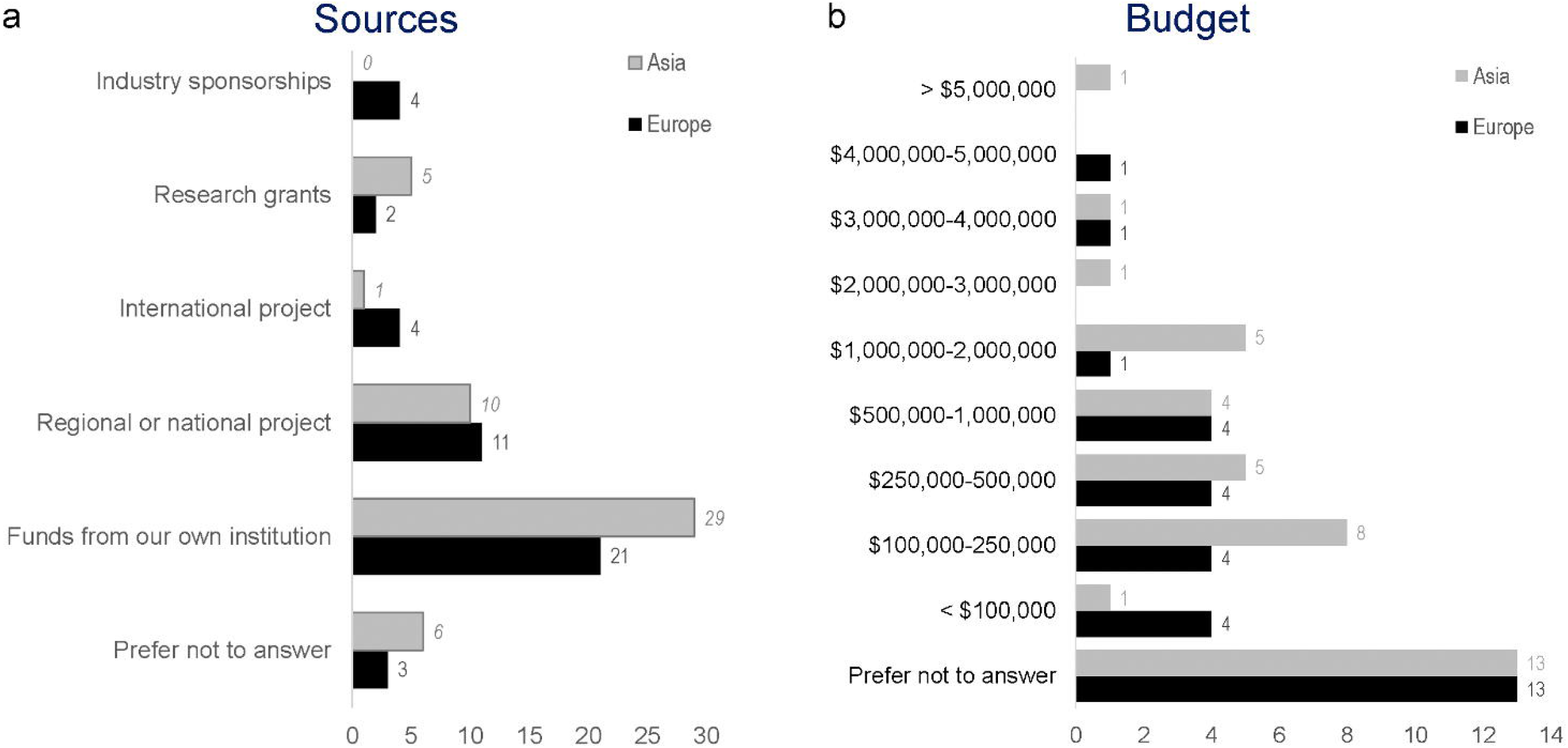
Expenses of the digital transition Costs per institution (**A**) and sources of funding (**B**)

### Software, including LIS and CAD algorithms

Digital pathology systems rely on a robust software backbone to enable fast and reliable access to WSIs, laboratory traceability, and the use of CAD tools. Integrating the system with the laboratory information system (LIS) is critical, as it allows for smoother and better-controlled workflows^7^.

Although most respondents claimed that they have some level of integration between the LIS and their WSI viewer (n = 41; 56.9%), many reported that no integration has been achieved, requiring them to open these two applications separately and manually (n = 28; 38.9%). The LIS was integrated with laboratory instruments in about two thirds of institutions (n = 45; 62.5%). Interestingly, WSI access was not limited to pathologists in most institutions (n = 48; 66.7%), with clinicians also having access to virtual slides for various purposes, usually via the local electronic medical records system.

Digital systems connected to the internet may allow remote access to institutional servers and workstations, facilitating telepathology. In this study, there was a nearly even split between laboratories that enabled remote access to WSIs and those that did not (n = 38; 52.8% vs. n = 34; 47.2%), with minimal differences between the European and Asian cohorts. However, this did not necessarily translate to pathologists being allowed to work from home, as only 26 (36.1%) institutions granted permission for remote sign-out. It should be noted that there was a statistically significant difference between Europe and Asia in this regard (p = 0.049), with working from home being much more common in European institutions compared to Asian ones (n = 18; 54.5% vs. n = 8; 20.5%). Additionally, 13 (18.1%) institutions emphasized that the COVID-19 pandemic accelerated the adoption of digital pathology due to the need for remote work during lockdowns, although the majority believed that the pandemic did not significantly impact their digital pathology implementation plans (n = 48; 66.7%). Technically, remote access was mostly facilitated through a VPN, either open-source or commercial (n = 40; 55.6%), usually with support from the informatics departments more commonly provided (n = 44; 61.1%).

Regarding the application of CAD, while a majority (n = 49; 68.1%) of respondents from digitized laboratories did not implement these tools, about a third (n = 23; 31.9%) had already done so. The most common use case of AI algorithms was the objective scoring/quantification of immunohistochemistry (n = 17; 23.6%). It’s worth noting that the overall perception of CAD and AI among pathologists was favorable, with a large majority (n = 63; 87.5%) stating that they believe in a synergy between AI and pathologists and that these tools would complement their current practice. A complete description of use cases, perceptions, and vendor choices is shown in Figure 4.

**Figure 4:**
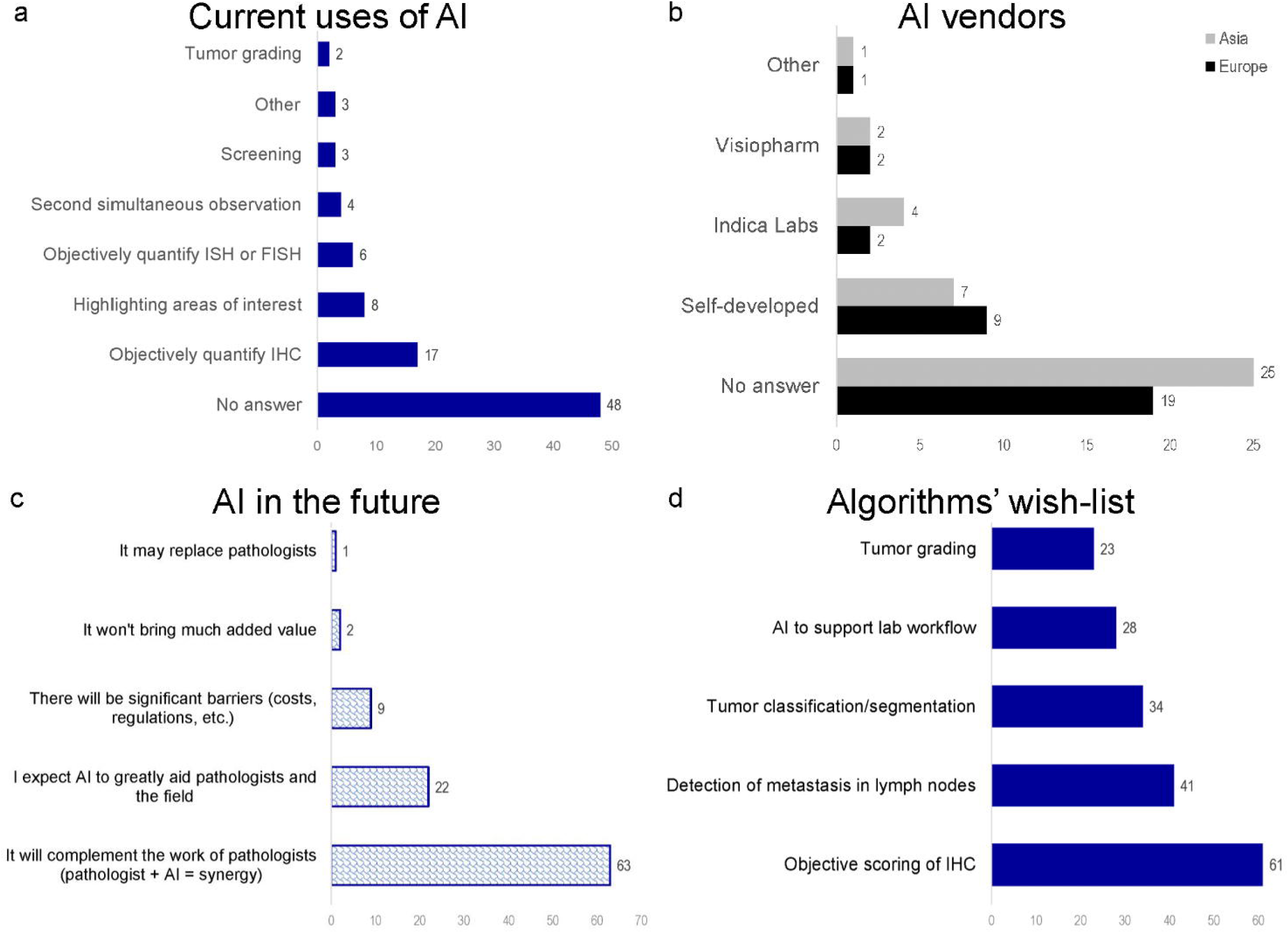
Adoption of AI by laboratories with digital workflow Current use cases (**A**), vendors (**B**), expectations for the future (**C**), and algorithms’ wish-list (**D**).

### Perceived benefits of digital pathology implementation

Overall, the outlooks about the impact of the implementation of digital pathology on the laboratory routine evaluated on a Likert scale from 1 (strongly disagree) to 5 (strongly agree) were positive, see details in Table 3. Most respondents agreed that it was useful for teaching and for tumor boards. Interestingly, respondents from Asia were more likely to find that digital pathology decreased response times for consultation cases compared to their European peers (median of 4.00 [IQR, 2.00] vs. 3.00 [IQR, 1.00]; p = 0.017). Quality control was seen as having improved. Most respondents declared that they wouldn’t go back to a microscope, with Asian pathologists being more likely to do so than their European peers (median of 2.00 [IQR, 2.00] vs. 1.00 [IQR, 1.00]; p = 0.03).

### Survey of laboratories without a digital pathology workflow

Out of 55 laboratories without access to digital pathology who participated in this survey, 43 (78.2%) were from Europe and 12 (21.8%) were from Asia. Responses came from 16 different countries, with 5 in Asia and 11 in Europe. Across all countries, the majority of respondents were from medium to small community hospitals (n = 24; 43.6%), followed by university hospitals (n = 19; 34.5%) and private laboratories (n = 12; 21.8%). Demographic details are shown in Figure 2. Most laboratories were staffed with either 1 to 5 (n = 26; 47.3%), 6 to 10 (n = 18; 32.7%), or 11 to 20 (n = 9; 16.4%) full-time pathologists, frequently supported by 1 to 5 part-time pathologists (n = 30; 54.5%). About half of the labs were involved in undergraduate medical education and postgraduate training in anatomic pathology.

Regarding previous experience with digital pathology, only 9 respondents (16.4%) stated they had never dealt with digital slides, while the rest had some experience either through online courses featuring virtual slides (n = 38; 69.1%), WSI repositories (n = 22; 40%), or accessing outside consultation cases (n = 6; 10.9%). In terms of anticipated future implementation, 36 respondents (65.5%) confirmed their plans to introduce digital pathology in the short or medium term, between 2 and 5 years.

Furthermore, most respondents considered lower costs of equipment and more available funding as possible incentives for installing scanners (n = 36; 65.5% and n = 28; 50.9%, respectively). Lower storage costs were also seen as important (n = 23; 41.8%), followed by easier access to expert opinions (n = 20; 36.4%), potential need for CAD according to upcoming international guidelines (n = 19; 34.5%), encouragement from hospital administration (n = 18; 32.7%), and faster scanning times (n = 17; 30.9%). Most respondents suggested that the adoption of digital pathology could be accelerated through the implementation of national or regional programs (n = 31; 56.4%).

Regarding obstacles, most respondents in this group were concerned about computer crashes (n = 17; 30.9%) and slow WSI loading times (n = 16; 29.1%), as well as the fact that scanning is an extra step in the laboratory workflow, which could lead to a longer turnaround time (n = 17; 30.9%). A significant proportion (n = 9; 16.4%) still believed that the standard microscope provided better diagnostic accuracy. Others were concerned that polarized light and cytology could not be digitized (n = 7; 12.7% each). Several respondents were worried about the possible unreliability of scan quality or the added complexity of scanning (n = 6; 10.9%). In addition, a significant proportion (n = 17; 30.9%) cited other reasons, mostly related to cost.

In terms of perceptions of digital pathology, nearly half of the respondents believed that the technology would transform the practice of anatomic pathology (n = 27; 49.1%). Only a small minority (n = 4; 7.3%) thought that the microscope would never be replaced. The last six questions of this section were answered on a Likert scale from 1 (strongly disagree) to 5 (strongly agree). The respondents showed high interest in adopting digital pathology in their institutions (median of 4.0 [IQR, 2.0]), but perceived cost as a major hindrance to adoption (median of 5.0 [IQR, 1.0]). Most agreed that there was a lack of appreciation for digital pathology among hospital/institution executives (median of 4.0 [IQR, 1.0]). Young pathologists were seen as drivers for its adoption (median of 4.0 [IQR, 1.0]), and most respondents did not see either pathologists or technicians as strong opponents of the digital transition (median of 2.0 [IQR, 2.0]).

## DISCUSSION

The implementation of digital pathology is not a simple endeavour and requires adjustments to the routine practice, including the laboratory workflow^7^. In our survey, we evaluated the process of digitization and current status in a significant number of institutions on two different continents, representing 69% of the world population and almost two-thirds of the global pathologist workforce^31,32^. Our data showed that, despite varying geographic, historical, social, and other factors, Europe and Asia did not differ significantly in most aspects of digital pathology implementation.

Digitizing a pathology laboratory is a new challenge, which just recently entered the stage, and according to our survey, most institutions began this process by scanning biopsies for teaching or tumor boards. At the same time, we observed a growing number of institutions with a more advanced level of digitization, including full or nearly full caseload scanning and the use of AI algorithms. Many other laboratories chose to focus only on certain subspecialties and digitize part of their routine cases. Given the trend toward adopting technology, these laboratories may still be in the early stages of the digital transition, with great potential to advance toward full digitization. Regarding the slide scanner, a centerpiece of the digital pathology laboratory, preference was given to medium- to high-throughput scanners, with limited use of single-slide digitizers and robotic microscopes.

Our data indicate that by the end of 2020 most institutions which adopted digital pathology workflow were large laboratories in university hospitals and cancer centers. A median of two workstations per laboratory suggests that they may not yet be equipped for the full-scale adoption of digital pathology. We believe that cost played a role as the main limiting factor. According to this survey, the average investment ranged from hundreds of thousands to millions of dollars, which could be prohibitive even for institutions with existing access to a scanner and even more so for those without one. The large storage needs may further exacerbate this problem.

Perceptions of potential concerns, such as increased turnaround time, computer crashes, or the perceived superiority of the microscope, may also interfere the full adoption of digital pathology. Based on our data, these concerns don’t seem to be entirely grounded in reality. While some issues persisted after implementation, increased turnaround time was not considered a major issue by the respondents. Slow loading times for virtual slides highlight the need for a good local area network connection and modern workstations, but the fact remains that most institutions that have implemented digital pathology would not return to a microscope-based routine. It is also concerning that many institutions did not employ validation protocols, did not report rescan rates, and lacked full sample traceability, which limits quality control and hinders further progress in workflow adaptations.

CAD algorithms were generally well received by both respondents with and without access to digital pathology but have not yet seen widespread adoption and use. Respondents indicated that these algorithms could be a major driver for future adoption of digital pathology, particularly, if they are incorporated into diagnostic schemes, reporting protocols, and clinical guidelines.

The differences between European and Asian laboratories are noteworthy. Pathologists from Asia were more likely to be concerned about delayed turnaround time. European laboratories that have adopted digital pathology stored WSIs for a shorter time than Asian labs, likely due to differences in legal frameworks. Asian institutions were more likely to have hired new employees to support the digital transition, such as technicians and secretaries, and were more likely to use smaller displays and less likely to allow working from home than European laboratories. Asian respondents felt that turnaround times for consultation cases improved with digital pathology, which was not acclaimed by European respondents.

Looking only at the cohort of pathologists without access to digital pathology, due to the pervasiveness of the technology, most respondents had encountered it either through online courses, WSI repositories, or consultation cases. More than half claimed to have plans for the implementation of digital pathology in some form in their laboratories in the short to medium term. Lower costs of equipment and more funding were seen the most critical factors to speed up adoption, highlighting once again the high costs of digital transition. National or regional strategic initiatives for the implementation could play a role in mitigating this factor.

This study has several limitations. Firstly, it provides only a single snapshot of the rapidly evolving field, which may become outdated within a few years. Secondly, the response rate to the email invitation was low, resulting in a lack of representation from some key countries, such as the United Kingdom. Additionally, institutions from the USA and Canada were deliberately excluded, as the authors are aware of several ongoing surveys focusing solely on this region. Despite these limitations, our study also has some strengths that are worth mentioning. This is the first and only large-scale, international survey on digital pathology implementation. It is comprehensive, covering a wide range of topics, from brands to staffing, and from tips to costs. Our survey, with its fully accessible data collection forms, can serve as a benchmark for comparison with future studies. The data presented here will be of interest to a diverse audience, including those in academia, industry, laboratory management, hospital administration, and policymaking.

In conclusion, while digital pathology is still the future for most, it is already the present for others, and there is no desire to come back after a digital transition. Our study is the first to describe this reality at an international scale. Digital pathology is well perceived by those who use it and those who do not. After implementation, the impact on the laboratory performance remains positive. CAD and AI, one of the major promises of digital pathology, saw limited use by the end of 2020. Costs are a major issue, but as technology advances and vendors compromise, we anticipate that equipment will become more affordable and interoperable. Nevertheless, regional, national, or international initiatives and funding programs would be of great help to drive digital pathology into mass adoption, which may be brought on by including AI algorithms in diagnostic and clinical guidelines. Unlike in the 1980s, when the first forays into digital pathology were made, today the digital transition is on solid ground. The 2020s might be the decade when pathology finally embraces the digital revolution.

## Data Availability

All data produced in the present work are contained in the manuscript

## STATEMENTS AND DECLARATIONS

## Acknowledgements

The authors would like to express their gratitude to all the participants who accepted the invitation to participate in this survey, for their valuable responses, feedback, and enthusiasm.

## Ethics Approval and Consent to Participate

Formal ethical approval was not required for this study, as it did not involve any patient data collection or impact on patient care. Participation was entirely voluntary, and there was no financial compensation for study participation and no disadvantage related to non-participation.

## Author Contributions

A.B. and C. E. performed study concept and design; D.G.P., A.B., and C. E. performed development of methodology and writing, review, and revision of the paper; A.B., N.T., J.F., and C.E. coordinated data collection; D.G.P. provided analysis and interpretation of data, and statistical analysis. All authors read and approved the final paper.

## Data Availability Statement

All data generated or analyzed during this study are included in this published article and its supplementary information files.

## SUPPLEMENTARY MATERIAL

**Supplementary Table 1**: Digital pathology implementation survey

**Supplementary Table 2**: Responses to sections 2–5 that targeted adopters of digital pathology only

**Supplementary Table 3**: Responses to section 6 addressed to non-adopters of digital pathology **Supplementary Table 4:** Survey items that reached statistically significant difference between European and Asian respondents

**Supplementary Table 5**: Comparison of responses from Europe and Asia

