## Supplemental tables 2-5 for "Exploring the adoption of digital pathology in clinical settings - Insights from a cross-continent study"

**SUPPLEMENTARY MATERIAL**

**Supplementary Table 2**: Responses to sections 2–5 that targeted adopters of digital pathology only

**Supplementary Table 3**: Responses to section 6 addressed to non-adopters of digital pathology

**Supplementary Table 4**: Survey items that reached statistically significant difference between European and Asian respondents

**Supplementary Table 5**: Comparison of responses from Europe and Asia

**Supplementary Table 2:** Responses to sections 2–5 that targeted adopters of digital pathology only

| Q | N | N % | Q | N | N % | Q | N | N % | Q | N | N % | Q | N | N % | Q | N | N % | Q | N | N % | Q | N | N % | Q | N | N % | Q | N | N % | Q | N | N % |
|---|---|---|---|---|---|---|---|---|---|---|---|---|---|---|---|---|---|---|---|---|---|---|---|---|---|---|---|---|---|---|---|---|
| **2.2** | | | **2.3** | | | **2.4** | | | **2.5.1** | | | **2.5.2** | | | **2.5.3** | | | **2.5.4** | | | **2.5.5** | | | **2.5.6** | | | **2.5.7** | | | **2.5.8** | | |
| **a)** | 12 | 16.7% | **a)** | 52 | 72.2% | **a)** | 68 | 94.4% | **a)** | 13 | 18.1% | **a)** | 12 | 16.7% | **a)** | 12 | 16.7% | **a)** | 12 | 16.7% | **a)** | 5 | 6.9% | **a)** | 12 | 16.7% | **a)** | 12 | 16.7% | **a)** | 14 | 19.4% |
| **b)** | 22 | 30.6% | **b)** | 55 | 76.4% | **b)** | 57 | 79.2% | **b)** | 42 | 58.3% | **b)** | 27 | 37.5% | **b)** | 30 | 41.7% | **b)** | 25 | 34.7% | **b)** | 5 | 6.9% | **b)** | 29 | 40.3% | **b)** | 30 | 41.7% | **b)** | 34 | 47.2% |
| **c)** | 15 | 20.8% | **c)** | 12 | 16.7% | **c)** | 17 | 23.6% | **c)** | 15 | 20.8% | **c)** | 28 | 38.9% | **c)** | 26 | 36.1% | **c)** | 24 | 33.3% | **c)** | 26 | 36.1% | **c)** | 24 | 33.3% | **c)** | 24 | 33.3% | **c)** | 20 | 27.8% |
| **d)** | 2 | 2.8% | **d)** | 27 | 37.5% | **d)** | 58 | 80.6% | **d)** | 2 | 2.8% | **d)** | 5 | 6.9% | **d)** | 4 | 5.6% | **d)** | 11 | 15.3% | **d)** | 36 | 50.0% | **d)** | 7 | 9.7% | **d)** | 6 | 8.3% | **d)** | 4 | 5.6% |
| **e)** | 3 | 4.2% | **e)** | 42 | 58.3% | **e)** | 58 | 80.6% | | | | | | | | | | | | | | | | | | | | | | | | |
| **f)** | 26 | 36.1% | **f)** | 18 | 25.0% | **f)** | 21 | 29.2% | | | | | | | | | | | | | | | | | | | | | | | | |
| **g)** | 43 | 59.7% | **g)** | 26 | 36.1% | **g)** | 1 | 1.4% | | | | | | | | | | | | | | | | | | | | | | | | |
| **h)** | 16 | 22.2% | **h)** | 51 | 70.8% | | | | | | | | | | | | | | | | | | | | | | | | | | | |
| **i)** | 2 | 2.8% | | | | | | | | | | | | | | | | | | | | | | | | | | | | | | |

| Q | N | N % | Q | N | N % | Q | N | N % | Q | N | N % | Q | N | N % | Q | N | N % | Q | N | N % | Q | N | N % | Q | N | N % | Q | N | N % | Q | N | N % |
|---|---|---|---|---|---|---|---|---|---|---|---|---|---|---|---|---|---|---|---|---|---|---|---|---|---|---|---|---|---|---|---|---|
| **2.5.9** | | | **2.5.10** | | | **2.5.11** | | | **2.5.12** | | | **2.5.13** | | | **2.5.14** | | | **2.5.15** | | | **2.5.16** | | | **2.6** | | | **2.7** | | | **2.8** | | |
| **a)** | 13 | 18.1% | **a)** | 12 | 16.7% | **a)** | 13 | 18.1% | **a)** | 11 | 15.3% | **a)** | 12 | 16.7% | **a)** | 12 | 16.7% | **a)** | 7 | 9.7% | **a)** | 5 | 6.9% | **a)** | 36 | 50.0% | **a)** | 41 | 56.9% | **a)** | 18 | 25.0% |
| **b)** | 34 | 47.2% | **b)** | 29 | 40.3% | **b)** | 18 | 25.0% | **b)** | 22 | 30.6% | **b)** | 24 | 33.3% | **b)** | 31 | 43.1% | **b)** | 12 | 16.7% | **b)** | 11 | 15.3% | **b)** | 13 | 18.1% | **b)** | 6 | 8.3% | **b)** | 21 | 29.2% |
| **c)** | 20 | 27.8% | **c)** | 26 | 36.1% | **c)** | 28 | 38.9% | **c)** | 27 | 37.5% | **c)** | 30 | 41.7% | **c)** | 25 | 34.7% | **c)** | 34 | 47.2% | **c)** | 46 | 63.9% | **c)** | 4 | 5.6% | **c)** | 10 | 13.9% | **c)** | 30 | 41.7% |
| **d)** | 5 | 6.9% | **d)** | 5 | 6.9% | **d)** | 13 | 18.1% | **d)** | 12 | 16.7% | **d)** | 6 | 8.3% | **d)** | 4 | 5.6% | **d)** | 19 | 26.4% | **d)** | 10 | 13.9% | **d)** | 3 | 4.2% | **d)** | 15 | 20.8% | **d)** | 23 | 31.9% |
| | | | | | | | | | | | | | | | | | | | | | | | | **e)** | 16 | 22.2% | | | | **e)** | 13 | 18.1% |
| | | | | | | | | | | | | | | | | | | | | | | | | | | | | | | **f)** | 17 | 23.6% |

| Q | N | N % |
|---|---|---|
| **2.9** a) | 36 | 50.0% |
| b) | 36 | 50.0% |

| Q | N | N % |
|---|---|---|
| **2.10** a) | 44 | 61.1% |
| b) | 16 | 22.2% |
| c) | 6 | 8.3% |
| d) | 0 | 0.0% |
| e) | 6 | 8.3% |

| Q | N | N % |
|---|---|---|
| **2.11** a) | 35 | 48.6% |
| b) | 34 | 47.2% |
| c) | 16 | 22.2% |
| d) | 7 | 9.7% |

| Q | N | N % |
|---|---|---|
| **2.12** a) | 49 | 68.1% |
| b) | 21 | 29.2% |
| c) | 2 | 2.8% |

| Q | N | N % |
|---|---|---|
| **2.14** a) | 33 | 45.8% |
| b) | 56 | 77.8% |
| c) | 22 | 30.6% |
| d) | 38 | 52.8% |
| e) | 29 | 40.3% |
| f) | 49 | 68.1% |
| g) | 42 | 58.3% |
| h) | 1 | 1.4% |

| Q | N | N % |
|---|---|---|
| **2.15** a) | 36 | 50.0% |
| b) | 32 | 44.4% |
| c) | 10 | 13.9% |
| d) | 9 | 12.5% |

| Q | N | N % |
|---|---|---|
| **2.16** a) | 14 | 19.4% |
| b) | 15 | 20.8% |
| c) | 14 | 19.4% |
| d) | 9 | 12.5% |
| e) | 20 | 27.8% |

| Q | N | N % |
|---|---|---|
| **2.17** a) | 44 | 61.1% |
| b) | 12 | 16.7% |
| c) | 13 | 18.1% |
| d) | 3 | 4.2% |

| Q | N | N % |
|---|---|---|
| **2.18** a) | 5 | 6.9% |
| b) | 12 | 16.7% |
| c) | 5 | 6.9% |
| d) | 9 | 12.5% |
| e) | 7 | 9.7% |
| f) | 6 | 8.3% |
| g) | 11 | 15.3% |
| h) | 47 | 65.3% |

| Q | N | N % |
|---|---|---|
| **2.19** a) | 25 | 34.7% |
| b) | 8 | 11.1% |
| c) | 17 | 23.6% |
| d) | 14 | 19.4% |
| e) | 15 | 20.8% |
| f) | 6 | 8.3% |
| g) | 23 | 31.9% |

| Q | N | N % |
|---|---|---|
| **2.20** a) | 29 | 40.3% |
| b) | 31 | 43.1% |
| c) | 42 | 58.3% |
| d) | 22 | 30.6% |
| e) | 1 | 1.4% |

| Q | N | N % |
|---|---|---|
| **2.21** a) | 41 | 56.9% |
| b) | 15 | 20.8% |
| c) | 4 | 5.6% |
| d) | 11 | 15.3% |
| e) | 4 | 5.6% |

| Q | N | N % |
|---|---|---|
| **2.22** a) | 41 | 56.9% |
| b) | 22 | 30.6% |
| c) | 12 | 16.7% |
| d) | 7 | 9.7% |
| e) | 3 | 4.2% |
| f) | 5 | 6.9% |
| g) | 1 | 1.4% |

| Q | N | N % |
|---|---|---|
| **2.23** a) | 30 | 41.7% |
| b) | 30 | 41.7% |
| c) | 47 | 65.3% |
| d) | 14 | 19.4% |
| e) | 10 | 13.9% |
| f) | 22 | 30.6% |

| Q | N | N % |
|---|---|---|
| **2.24** a) | 36 | 50.0% |
| b) | 35 | 48.6% |
| c) | 1 | 1.4% |

| Q | N | N % |
|---|---|---|
| **2.25** a) | 10 | 13.9% |
| b) | 55 | 76.4% |
| c) | 7 | 9.7% |
| d) | 0 | 0.0% |

| Q | N | N % |
|---|---|---|
| **2.26** a) | 35 | 48.6% |
| b) | 36 | 50.0% |
| c) | 19 | 26.4% |
| d) | 7 | 9.7% |
| e) | 38 | 52.8% |
| f) | 32 | 44.4% |
| g) | 7 | 9.7% |

| Q | N | N % |
|---|---|---|
| **3.1** a) | 37 | 51.4% |
| b) | 18 | 25.0% |
| c) | 6 | 8.3% |
| d) | 6 | 8.3% |
| e) | 1 | 1.4% |
| f) | 4 | 5.6% |

| Q | N | N % |
|---|---|---|
| **3.2** a) | 10 | 13.9% |
| b) | 16 | 22.2% |
| c) | 15 | 20.8% |
| d) | 14 | 19.4% |
| e) | 25 | 34.7% |
| f) | 3 | 4.2% |

| Q | N | N % |
|---|---|---|
| **3.3** a) | 8 | 11.1% |
| b) | 9 | 12.5% |
| c) | 11 | 15.3% |
| d) | 2 | 2.8% |
| e) | 42 | 59.7% |
| f) | 2 | 2.8% |

| Q | N | N % |
|---|---|---|
| **3.4** a) | 3 | 4.2% |
| b) | 13 | 18.1% |
| c) | 3 | 4.2% |
| d) | 4 | 5.6% |
| e) | 43 | 59.7% |
| f) | 9 | 12.5% |

| Q | N | N % |
|---|---|---|
| **3.5** | | |
| **a)** | 64 | 88.9% |
| **b)** | 52 | 72.2% |
| **c)** | 45 | 62.5% |
| **d)** | 51 | 70.8% |
| **e)** | 14 | 19.4% |
| **f)** | 13 | 18.1% |
| **g)** | 4 | 5.6% |
| **h)** | 22 | 30.6% |
| **i)** | 39 | 54.2% |
| **j)** | 38 | 52.8% |
| **k)** | 1 | 1.4% |

| Q | N | N % |
|---|---|---|
| **3.6** | | |
| **a)** | 6 | 8.3% |
| **b)** | 14 | 19.4% |
| **c)** | 39 | 54.2% |
| **d)** | 13 | 18.1% |
| **e)** | 0 | 0.0% |

| Q | N | N % |
|---|---|---|
| **3.7** | | |
| **a)** | 48 | 66.7% |
| **b)** | 23 | 31.9% |
| **c)** | 1 | 1.4% |

| Q | N | N % |
|---|---|---|
| **3.8** | | |
| **a)** | 58 | 80.6% |
| **b)** | 7 | 9.7% |
| **c)** | 6 | 8.3% |
| **d)** | 1 | 1.4% |

| Q | N | N % |
|---|---|---|
| **3.9** | | |
| **a)** | 24 | 33.3% |
| **b)** | 7 | 9.7% |
| **c)** | 41 | 56.9% |

| Q | N | N % |
|---|---|---|
| **3.10** | | |
| **a)** | 26 | 36.1% |
| **b)** | 5 | 6.9% |
| **c)** | 17 | 23.6% |
| **d)** | 24 | 33.3% |

| Q | N | N % |
|---|---|---|
| **3.11** | | |
| **a)** | 46 | 63.9% |
| **b)** | 12 | 16.7% |
| **c)** | 14 | 19.4% |

| Q | N | N % |
|---|---|---|
| **3.12** | | |
| **a)** | 65 | 90.3% |
| **b)** | 3 | 4.2% |
| **c)** | 4 | 5.6% |

| Q | N | N % |
|---|---|---|
| **3.14** | | |
| **a)** | 12 | 16.7% |
| **b)** | 29 | 40.3% |
| **c)** | 20 | 27.8% |
| **d)** | 9 | 12.5% |
| **e)** | 2 | 2.8% |

| Q | N | N % |
|---|---|---|
| **3.15** | | |
| **a)** | 4 | 5.6% |
| **b)** | 8 | 11.1% |
| **c)** | 49 | 68.1% |
| **d)** | 6 | 8.3% |
| **e)** | 5 | 6.9% |

| Q | N | N % |
|---|---|---|
| **3.16** | | |
| **a)** | 17 | 23.6% |
| **b)** | 13 | 18.1% |
| **c)** | 7 | 9.7% |
| **d)** | 7 | 9.7% |
| **e)** | 8 | 11.1% |
| **f)** | 20 | 27.8% |

| Q | N | N % |
|---|---|---|
| **3.17** | 38 | 52.8% |
| **a)** | | |
| **b)** | 19 | 26.4% |
| **c)** | 7 | 9.7% |
| **d)** | 8 | 11.1% |

| Q | N | N % |
|---|---|---|
| **3.19** | 47 | 65.3% |
| **a)** | | |
| **b)** | 1 | 1.4% |
| **c)** | 11 | 15.3% |
| **d)** | 3 | 4.2% |
| **e)** | 8 | 11.1% |
| **f)** | 2 | 2.8% |

| Q | N | N % |
|---|---|---|
| **3.20** | 44 | 61.1% |
| **a)** | | |
| **b)** | 28 | 38.9% |

| Q | N | N % |
|---|---|---|
| **3.21** | 9 | 12.5% |
| **a)** | | |
| **b)** | 6 | 8.3% |
| **c)** | 17 | 23.6% |
| **d)** | 27 | 37.5% |
| **e)** | 13 | 18.1% |

| Q | N | N % |
|---|---|---|
| **3.22** | 33 | 45.8% |
| **a)** | | |
| **b)** | 39 | 54.2% |
| **c)** | 0 | 0.0% |

| Q | N | N % |
|---|---|---|
| **3.26** | 34 | 47.2% |
| **a)** | | |
| **b)** | 17 | 23.6% |
| **c)** | 8 | 11.1% |

| Q | N | N % |
|---|---|---|
| **3.28** | 5 | 6.9% |
| **a)** | | |
| **b)** | 27 | 37.5% |
| **c)** | 10 | 13.9% |
| **d)** | 1 | 1.4% |
| **e)** | 17 | 23.6% |
| **f)** | 2 | 2.8% |
| **g)** | 10 | 13.9% |

| Q | N | N % |
|---|---|---|
| **3.29** | 4 | 5.6% |
| **a)** | | |
| **b)** | 1 | 1.4% |
| **c)** | 17 | 23.6% |
| **d)** | 5 | 6.9% |
| **e)** | 45 | 62.5% |
| **f)** | 0 | 0.0% |

| Q | N | N % |
|---|---|---|
| **3.30** | 12 | 16.7% |
| **a)** | | |
| **b)** | 2 | 2.8% |
| **c)** | 9 | 12.5% |
| **d)** | 23 | 31.9% |
| **e)** | 26 | 36.1% |
| **f)** | 0 | 0.0% |

| Q | N | N % |
|---|---|---|
| **3.31** | 42 | 58.3% |
| **a)** | | |
| **b)** | 26 | 36.1% |
| **c)** | 4 | 5.6% |

| Q | N | N % |
|---|---|---|
| **3.32** | 39 | 54.2% |
| **a)** | | |
| **b)** | 44 | 61.1% |
| **c)** | 60 | 83.3% |
| **d)** | 14 | 19.4% |
| **e)** | 35 | 48.6% |
| **f)** | 7 | 9.7% |
| **g)** | 5 | 6.9% |

| Q | N | N % |
|---|---|---|
| **3.33** | | |
| **a)** | 26 | 36.1% |
| **b)** | 5 | 6.9% |
| **c)** | 12 | 16.7% |
| **d)** | 9 | 12.5% |
| **e)** | 8 | 11.1% |

| Q | N | N % |
|---|---|---|
| **3.34** | | |
| **a)** | 9 | 12.5% |
| **b)** | 50 | 69.4% |
| **c)** | 21 | 29.2% |
| **d)** | 5 | 6.9% |
| **e)** | 7 | 9.7% |

| Q | N | N % |
|---|---|---|
| **3.35** | | |
| **a)** | 14 | 19.4% |
| **b)** | 12 | 16.7% |
| **c)** | 15 | 20.8% |
| **d)** | 31 | 43.1% |
| **e)** | 0 | 0.0% |

| Q | N | N % |
|---|---|---|
| **4.1** | | |
| **a)** | 28 | 38.9% |
| **b)** | 9 | 12.5% |
| **c)** | 16 | 22.2% |
| **d)** | 16 | 22.2% |
| **e)** | 3 | 4.2% |

| Q | N | N % |
|---|---|---|
| **4.2** | | |
| **a)** | 45 | 62.5% |
| **b)** | 27 | 37.5% |
| **c)** | 0 | 0.0% |

| Q | N | N % |
|---|---|---|
| **4.3** | | |
| **a)** | 38 | 52.8% |
| **b)** | 34 | 47.2% |
| **c)** | 0 | 0.0% |

| Q | N | N % |
|---|---|---|
| **4.4** | | |
| **a)** | 25 | 34.7% |
| **b)** | 15 | 20.8% |
| **c)** | 6 | 8.3% |
| **d)** | 5 | 6.9% |
| **e)** | 1 | 1.4% |

| Q | N | N % |
|---|---|---|
| **4.5** | | |
| **a)** | 9 | 12.5% |
| **b)** | 35 | 48.6% |
| **c)** | 5 | 6.9% |
| **d)** | 0 | 0.0% |
| **e)** | 0 | 0.0% |

| Q | N | N % |
|---|---|---|
| **4.6** | | |
| **a)** | 26 | 36.1% |
| **b)** | 46 | 63.9% |
| **c)** | 0 | 0.0% |

| Q | N | N % |
|---|---|---|
| **4.7** | 26 | 36.1% |
| **a)** | | |
| **b)** | 43 | 59.7% |
| **c)** | 3 | 4.2% |

| Q | N | N % |
|---|---|---|
| **4.8** | | |
| **a)** | 0 | 0.0% |
| **b)** | 58 | 80.6% |
| **c)** | 3 | 4.2% |
| **d)** | 3 | 4.2% |
| **e)** | 16 | 22.2% |

| | | | | | | | | | | | | | | | | | | | | | | | | | | | | | | | | |
|---|---|---|---|---|---|---|---|---|---|---|---|---|---|---|---|---|---|---|---|---|---|---|---|---|---|---|---|---|---|---|---|---|
| **f)** | 6 | 8.3% | **f)** | 4 | 5.6% | | | | | | | | | | | | | **f)** | 32 | 44.4% | **f)** | 9 | 12.5% | | | | | | | **f)** | 11 | 15.3% |
| **g)** | 1 | 1.4% | **g)** | 0 | 0.0% | | | | | | | | | | | | | | | | | | | | | | | | | **g)** | 8 | 11.1% |
| **h)** | 2 | 2.8% | | | | | | | | | | | | | | | | | | | | | | | | | | | | | | **h)** | 7 | 9.7% |
| **i)** | 1 | 1.4% | | | | | | | | | | | | | | | | | | | | | | | | | | | | | | **i)** | 0 | 0.0% |
| **j)** | 1 | 1.4% | | | | | | | | | | | | | | | | | | | | | | | | | | | | | | | | |
| **k)** | 1 | 1.4% | | | | | | | | | | | | | | | | | | | | | | | | | | | | | | | | |

| Q | N | N % |
|---|---|---|
| **4.9** a) | 13 | 18.1% |
| b) | 10 | 13.9% |
| c) | 48 | 66.7% |
| d) | 1 | 1.4% |

| Q | N | N % |
|---|---|---|
| **4.10** a) | 23 | 31.9% |
| b) | 49 | 68.1% |
| c) | 0 | 0.0% |

| Q | N | N % |
|---|---|---|
| **4.11** a) | 2 | 2.8% |
| b) | 17 | 23.6% |
| c) | 6 | 8.3% |
| d) | 8 | 11.1% |
| e) | 3 | 4.2% |
| f) | 4 | 5.6% |
| g) | 3 | 4.2% |
| h) | 48 | 66.7% |

| Q | N | N % |
|---|---|---|
| **4.12** a) | 16 | 22.2% |
| b) | 0 | 0.0% |
| c) | 0 | 0.0% |
| d) | 4 | 5.6% |
| e) | 6 | 8.3% |
| f) | 0 | 0.0% |
| g) | 2 | 2.8% |
| h) | 44 | 61.1% |

| Q | N | N % |
|---|---|---|
| **4.13** a) | 1 | 1.4% |
| b) | 63 | 87.5% |
| c) | 2 | 2.8% |
| d) | 9 | 12.5% |
| e) | 22 | 30.6% |
| f) | 0 | 0.0% |

| Q | N | N % |
|---|---|---|
| **4.14** a) | 61 | 84.7% |
| b) | 23 | 31.9% |
| c) | 34 | 47.2% |
| d) | 41 | 56.9% |
| e) | 28 | 38.9% |
| f) | 0 | 0.0% |

| Q | N | N % |
|---|---|---|
| **5.1** 1) | 12 | 16.7% |
| 2) | 19 | 26.4% |
| 3) | 24 | 33.3% |
| 4) | 14 | 19.4% |
| 5) | 3 | 4.2% |
| x̄ | 2.68 | |
| σ | 1.10 | |

| Q | N | N % |
|---|---|---|
| **5.2** 1) | 7 | 9.7% |
| 2) | 5 | 6.9% |
| 3) | 22 | 30.6% |
| 4) | 17 | 23.6% |
| 5) | 21 | 29.2% |
| x̄ | 3.56 | |
| σ | 1.25 | |

| Q | N | N % |
|---|---|---|
| **5.3** 1) | 4 | 5.6% |
| 2) | 3 | 4.2% |
| 3) | 24 | 33.3% |
| 4) | 24 | 33.3% |
| 5) | 17 | 23.6% |
| x̄ | 3.65 | |
| σ | 1.06 | |

| Q | N | N % |
|---|---|---|
| **5.4** 1) | 5 | 6.9% |
| 2) | 6 | 8.3% |
| 3) | 22 | 30.6% |
| 4) | 17 | 23.6% |
| 5) | 22 | 30.6% |
| x̄ | 3.63 | |
| σ | 1.20 | |

| Q | N | N % |
|---|---|---|
| **5.5** 1) | 1 | 1.4% |
| 2) | 0 | 0.0% |
| 3) | 4 | 5.6% |
| 4) | 20 | 27.8% |
| 5) | 47 | 65.3% |
| x̄ | 4.56 | |
| σ | 0.73 | |

| Q | N | N % |
|---|---|---|
| **5.6** 1) | 2 | 2.8% |
| 2) | 1 | 1.4% |
| 3) | 5 | 6.9% |
| 4) | 18 | 25.0% |
| 5) | 46 | 63.9% |
| x̄ | 4.46 | |
| σ | 0.90 | |

| Q | N | N % |
|---|---|---|
| **5.7** 1) | 10 | 13.9% |
| 2) | 9 | 12.5% |
| 3) | 21 | 29.2% |
| 4) | 15 | 20.8% |
| 5) | 17 | 23.6% |
| x̄ | 3.29 | |
| σ | 1.33 | |

| Q | N | N % |
|---|---|---|
| **5.8** 1) | 3 | 4.2% |
| 2) | 7 | 9.7% |
| 3) | 17 | 23.6% |
| 4) | 28 | 38.9% |
| 5) | 17 | 23.6% |
| x̄ | 3.68 | |
| σ | 1.07 | |

| Q | N | N % |
|---|---|---|
| **5.9** 1) | 3 | 4.2% |
| 2) | 9 | 12.5% |
| 3) | 32 | 44.4% |
| 4) | 20 | 27.8% |
| 5) | 8 | 11.1% |
| x̄ | 3.29 | |
| σ | 0.97 | |

| Q | N | N % |
|---|---|---|
| **5.10** 1) | 9 | 12.5% |
| 2) | 12 | 16.7% |
| 3) | 14 | 19.4% |
| 4) | 26 | 36.1% |
| 5) | 11 | 15.3% |
| x̄ | 3.25 | |
| σ | 1.264 | |

| Q | N | N % |
|---|---|---|
| **5.11** 1) | 15 | 20.8% |
| 2) | 14 | 19.4% |
| 3) | 20 | 27.8% |
| 4) | 17 | 23.6% |
| 5) | 6 | 8.3% |
| x̄ | 2.79 | |
| σ | 1.26 | |

| Q | N | N % |
|---|---|---|
| **5.12** 1) | 8 | 11.1% |
| 2) | 14 | 19.4% |
| 3) | 29 | 40.3% |
| 4) | 16 | 22.2% |
| 5) | 5 | 6.9% |
| x̄ | 2.94 | |
| σ | 1.07 | |

| Q | N | N % |
|---|---|---|
| **5.13** 1) | 6 | 8.3% |
| 2) | 9 | 12.5% |
| 3) | 19 | 26.4% |
| 4) | 28 | 38.9% |
| 5) | 10 | 13.9% |
| x̄ | 3.38 | |
| σ | 1.13 | |

| Q | N | N % |
|---|---|---|
| **5.14** 1) | 16 | 22.2% |
| 2) | 13 | 18.1% |
| 3) | 26 | 36.1% |
| 4) | 12 | 16.7% |
| 5) | 5 | 6.9% |
| x̄ | 2.68 | |
| σ | 1.20 | |

| Q | N | N % |
|---|---|---|
| **5.15** 1) | 40 | 55.6% |
| 2) | 14 | 19.4% |
| 3) | 14 | 19.4% |
| 4) | 1 | 1.4% |
| 5) | 3 | 4.2% |
| x̄ | 1.79 | |
| σ | 1.07 | |

| Q | N | N % |
|---|---|---|
| **5.16** 1) | 6 | 8.3% |
| 2) | 8 | 11.1% |
| 3) | 15 | 20.8% |
| 4) | 27 | 37.5% |
| 5) | 16 | 22.2% |
| x̄ | 3.54 | |
| σ | 1.19 | |

**Supplementary Table 3:** Responses to section 6 addressed to non-adopters of digital pathology

| Q | N | N % | Q | N | N % | Q | N | N % | Q | N | N % | Q | N | N % | Q | N | N % | Q | N | N % | Q | N | N % | Q | N | N % | Q | N | N % | Q | N | N % |
|---|---|---|---|---|---|---|---|---|---|---|---|---|---|---|---|---|---|---|---|---|---|---|---|---|---|---|---|---|---|---|---|---|
| <u>**6.1**</u><br>**a)** | 9 | 16.4% | <u>**6.2**</u><br>**a)** | 20 | 36.4% | <u>**6.3**</u><br>**a)** | 36 | 65.5% | <u>**6.4**</u><br>**a)** | 17 | 30.9% | <u>**6.5**</u><br>**a)** | 4 | 7.3% | <u>**6.6**</u><br>**1)** | 6 | 10.9% | <u>**6.7**</u><br>**1)** | 0 | 0.0% | <u>**6.8**</u><br>**1)** | 7 | 12.7% | <u>**6.9**</u><br>**1)** | 11 | 20.0% | <u>**6.10**</u><br>**1)** | 26 | 47.3% | <u>**6.11**</u><br>**1)** | 16 | 29.1% |
| **b)** | 38 | 69.1% | **b)** | 16 | 29.1% | **b)** | 23 | 41.8% | **b)** | 16 | 29.1% | **b)** | 14 | 25.5% | **2)** | 7 | 12.7% | **2)** | 5 | 9.1% | **2)** | 4 | 7.3% | **2)** | 2 | 3.6% | **2)** | 9 | 16.4% | **2)** | 12 | 21.8% |
| **c)** | 22 | 40.0% | **c)** | 7 | 12.7% | **c)** | 17 | 30.9% | **c)** | 6 | 10.9% | **c)** | 5 | 9.1% | **3)** | 7 | 12.7% | **3)** | 8 | 14.5% | **3)** | 17 | 30.9% | **3)** | 14 | 25.5% | **3)** | 9 | 16.4% | **3)** | 15 | 27.3% |
| **d)** | 6 | 10.9% | **d)** | 10 | 18.2% | **d)** | 19 | 34.5% | **d)** | 6 | 10.9% | **d)** | 5 | 9.1% | **4)** | 16 | 29.1% | **4)** | 10 | 18.2% | **4)** | 13 | 23.6% | **4)** | 17 | 30.9% | **4)** | 3 | 5.5% | **4)** | 7 | 12.7% |
| **e)** | 11 | 20.0% | **e)** | 2 | 3.6% | **e)** | 20 | 36.4% | **e)** | 7 | 12.7% | **e)** | 27 | 49.1% | **5)** | 19 | 34.5% | **5)** | 32 | 58.2% | **5)** | 14 | 25.5% | **5)** | 11 | 20.0% | **5)** | 8 | 14.5% | **5)** | 5 | 9.1% |
| | | | | | | **f)** | 28 | 50.9% | **f)** | 7 | 12.7% | | | | $\bar{x}$ | 3.63 | | $\bar{x}$ | 4.25 | | $\bar{x}$ | 3.42 | | $\bar{x}$ | 3.27 | | $\bar{x}$ | 2.23 | | $\bar{x}$ | 2.51 | |
| | | | | | | **g)** | 11 | 20.0% | **g)** | 9 | 16.4% | | | | $\sigma$ | 1.37 | | $\sigma$ | 1.02 | | $\sigma$ | 1.3 | | $\sigma$ | 1.38 | | $\sigma$ | 1.47 | | $\sigma$ | 1.29 | |
| | | | | | | **h)** | 18 | 32.7% | **h)** | 17 | 30.9% | | | | | | | | | | | | | | | | | | | | | |
| | | | | | | **i)** | 7 | 12.7% | **i)** | 17 | 30.9% | | | | | | | | | | | | | | | | | | | | | |
| | | | | | | **j)** | 31 | 56.4% | | | | | | | | | | | | | | | | | | | | | | | | |
| | | | | | | **k)** | 0 | 0.0% | | | | | | | | | | | | | | | | | | | | | | | | |

**Supplementary Table 4:** Survey items that reached statistically significant difference between European and Asian respondents

| | | | | | |
|---|---|---|---|---|---|
| **Question 2.8** | If there are pathologists that do not work with digital pathology in your institution, can you elaborate on the reasons why? | | | | |
| Multiple choice, multiple answers | **Europe** | **%** | **Asia** | **%** | **p-value** |
| Limited access to workstations | 6 | 18.2% | 12 | 30.8% | **0.0010*** |
| Some pathologists aren't happy with a quality of digital slides | 3 | 9.1% | 18 | 46.2% | |
| Some pathologists are personally/internally against digital pathology | 10 | 30.3% | 20 | 51.3% | |
| Due to slower turn-around-time of digital workflow | 4 | 12.1% | 19 | 48.7% | |
| Some pathologists are specialized in areas that aren't digitized | 8 | 24.2% | 5 | 12.8% | |
| No response | 12 | 36.4% | 5 | 12.8% | |
| **Question 2.22** | Did you need to hire new personnel during the process of implementing a digital pathology workflow? | | | | |
| Multiple choice, multiple answers | **Europe** | **%** | **Asia** | **%** | **p-value** |
| No | 26 | 78.8% | 15 | 38.5% | |
| Technicians / Biomedical scientists | 5 | 15.2% | 17 | 43.6% | **0.0065*** |
| Secretaries | 1 | 3.0% | 11 | 28.2% | |
| Pathologists (to supervise/ administrate digital pathology, e.g. Head of Digital Pathology) | 3 | 9.1% | 4 | 10.3% | |
| Pathologists (for diagnostic purposes) | 2 | 6.1% | 1 | 2.6% | |
| IT personnel | 3 | 9.1% | 2 | 5.1% | |
| Other (short text) | 1 | 3.0% | 0 | 0.0% | |
| **Question 3.15** | What is your current storage plan/solution for digital slides? | | | | |
| Multiple choice, multiple answers | **Europe** | **%** | **Asia** | **%** | **p-value** |
| Digital slides are quickly discarded (in a few months) | 4 | 12.1% | 0 | 0.0% | **0.0006*** |
| Digital slides are stored for a short period (< 1 year) | 7 | 21.2% | 1 | 2.6% | |
| Digital slides are stored for a long period (≥ 1 year) | 17 | 51.5% | 32 | 82.1% | |
| I don't know | 1 | 3.0% | 5 | 12.8% | |
| Other (short text) | 4 | 12.1% | 1 | 2.6% | |
| **Question 3.25** | What is the standard screen size (for WSI viewing) of your workstation? (in inches) | | | | |
| Short text (open-ended) | **Europe** | | **Asia** | | **p-value** |
| Median – Interquartile interval | 27.0 | 7.3 | 25.0 | 3.0 | **0.0234*** |
| Minimum – Maximum | 20 | 51 | 17 | 48 | |
| **Question 4.7** | Are pathologists allowed to diagnose/work from home at your institution? | | | | |
| Multiple choice, single answer | **Europe** | **%** | **Asia** | **%** | **p-value** |
| Yes | 18 | 54.5% | 8 | 20.5% | **0.0492*** |
| No | 12 | 36.4% | 31 | 79.5% | |
| **Question 5.4** | The implementation of digital pathology has improved the response time of consultation cases | | | | |
| Likert Scale, 1 (strongly disagree) to 5 (strongly agree) | **Europe** | | **Asia** | | **p-value** |
| Median – Interquartile interval | 3.0 | 1.0 | 4.0 | 2.0 | **0.0017*** |
| Minimum – Maximum | 1 | 5 | 1 | 5 | |
| **Question 5.15** | If I had the option, I would step back to classical/analogue microscopy and significantly limit use of digital pathology | | | | |
| Likert Scale, 1 (strongly disagree) to 5 (strongly agree) | **Europe** | | **Asia** | | **p-value** |
| Median – Interquartile interval | 1.0 | 1.0 | 2.0 | 2.0 | **0.0301*** |
| Minimum – Maximum | 1 | 3 | 1 | 5 | |

**Supplementary Table 5:** Comparison of responses from Europe and Asia

| Question | p-value | Question | p-value | Question | p-value | Question | p-value |
|---|---|---|---|---|---|---|---|
| **2.1** | 0.0665 | **2.17** | 0.3346 | **3.22** | 0.2359 | **5.4** | **0,017*** |
| **2.2** | 0.8836 | **2.18** | 0.4266[1] | **3.23** | 0,0885 | **5.5** | 0,5092 |
| **2.3** | 0.05587 | **2.19** | 0.9842 | **3.24** | 0,3051 | **5.6** | 0,191 |
| **2.4** | 0.9171 | **2.20** | 0.9769 | **3.25** | **0,0234*** | **5.7** | 0,6704 |
| **2.5.1** | 0.4077 | **2.21** | 0.6314 | **3.26** | 0.4348 | **5.8** | 0,3502 |
| **2.5.2** | >0.9999 | **2.22** | **0.0065*** | **3.27** | 0.5308[1] | **5.9** | 0,8986 |
| **2.5.3** | 0.8121 | **2.23** | 0.8930 | **3.28** | 0.8039 | **5.10** | 0,2317 |
| **2.5.4** | 0.6442 | **2.24** | 0.6365 | **3.29** | >0.9999 | **5.11** | 0,3261 |
| **2.5.5** | 0.0970 | **2.25** | >0.9999 | **3.30** | 0.4483 | **5.12** | 0,6819 |
| **2.5.6** | >0.9999 | **2.26** | 0.4728 | **3.31** | >0.9999 | **5.13** | 0,732 |
| **2.5.7** | 0.4754 | **3.1** | 0.2708 | **3.32** | 0.7239 | **5.14** | 0,0546 |
| **2.5.8** | 0.8023 | **3.2** | 0.0925[1] | **3.33** | 0.5593 | **5.15** | **0,0301*** |
| **2.5.9** | >0.9999 | **3.3** | 0.2683 | **3.34** | 0.3284 | **5.16** | 0,6595 |
| **2.5.10** | 0.6366 | **3.4** | 0.1041 | **3.35** | 0.3400 | **6.1** | 0.2580 |
| **2.5.11** | 0.8123 | **3.5** | 0.8649 | **4.1** | 0.2073 | **6.2** | 0.3279 |
| **2.5.12** | 0.6407 | **3.6** | 0.7835 | **4.2** | 0.6263 | **6.3** | 0.7950[1] |
| **2.5.13** | 0.6365 | **3.7** | 0.8023 | **4.3** | 0.1032 | **6.4** | 0.9414[1] |
| **2.5.14** | 0.3375 | **3.8** | 0.3579 | **4.4** | 0.5077 | **6.5** | 0.9316[1] |
| **2.5.15** | 0.4276 | **3.9** | 0.1435 | **4.5** | >0.9999 | **6.6** | 0.9563 |
| **2.5.16** | >0.9999 | **3.10** | 0.3175 | **4.6** | 0.3345 | **6.7** | 0.3788 |
| **2.6** | >0.9999 | **3.11** | 0.3345 | **4.7** | **0.0492*** | **6.8** | 0.7111 |
| **2.7** | 0.4675 | **3.12** | 0.6954 | **4.8** | 0.0463 | **6.9** | 0.7382 |
| **2.8** | **0.0010*** | **3.13** | 0.6152 | **4.9** | 0.3890 | **6.10** | 0.3566 |
| **2.9** | 0.1554 | **3.14** | 0.7535 | **4.10** | 0.1272 | **6.11** | 0.3617 |
| **2.10** | 0.9056 | **3.15** | **0.0006*** | **4.11** | 0.4527 | | |
| **2.11** | 0.1022 | **3.16** | 0.6505 | **4.12** | 0.7345 | | |
| **2.12** | 0.1156 | **3.17** | >0.9999 | **4.13** | >0.9999 | | |
| **2.13** | >0.9999 | **3.18 (D/U)** | 0,9638 / 0.0988 | **4.14** | 0.2662 | | |
| **2.14** | 0.8956 | **3.19** | 0.3070 | **5.1** | 0.7245 | | |
| **2.15** | 0.1364 | **3.20** | 0.8093 | **5.2** | 0.3221 | | |
| **2.16** | 0.4849 | **3.21** | 0.0797 | **5.3** | 0.2485 | | |

[1] Chi-square requirements not met
